# The energy balance theory: an unsatisfactory model of body composition fluctuations

**DOI:** 10.1101/2020.10.27.20220202

**Authors:** Francisco Arencibia-Albite, Anssi H. Manninen

## Abstract

Differential weight and fat losses under isocaloric diets of distinct macronutrient composition are well-documented findings in obesity research.^1-6^ Such data are considered a result of inadequate methodology as it disagree with the energy balance theory.^7^ A recent mathematical analysis of this paradigm has found, however, serious analytical contradictions in its foundations.^8^ As an alternative, a mass balance model was proposed to explain the aforesaid body composition alterations. Here, we expand on this observation by contrasting both models. We show that mass balance explains a wide range of fending experiments including those concurring with the energy balance principle. The latter, however, is less flexible and results in poor forecasts in settings consistent with mass balance. The energy balance theory is thus an unsatisfactory model of body composition changes. Consequently, by shifting to a mass balance paradigm of obesity a much deeper understanding of this disease may follow in the near future.

## Introduction

The energy balance theory (EBT) is fundamentally a descriptive theory and only a small number of researchers had offer a formal quantitative framework of its essential principles.^9-13^ These treatments had been developed under the assumption that the all the EBT’s axioms are valid statements. Arencibia-Albite^8^ has recently performed a mathematical analysis of the EBT that demonstrates that the central thesis of theory is logically inconsistent. He showed, in particular, that body weight stability will always coexists with a persistent energy imbalanced. Such analytical result do not represent a violation of the First Law of Thermodynamics, as this principle allows for any open system to express a positive or negative energy balance under a null mass change.^8^

As an alternative to the EBT, a mass balance model (MBM) was proposed.^8^ It maintains that *body weight fluctuations are dependent on the difference between daily mass intake, in food and beverages, and daily mass excretion* (e.g., elimination of macronutrient oxidation products) and not on energy imbalance. This model fitted weight loss data from dietary interventions of low-carbohydrate diets (LCDs) versus isocaloric low-fat diets (LFDs) leading to a parsimonious account for the differential weigh and fat losses not explained by standard EBT-based hypothesis; i.e., energy underreporting, energy imbalance, glycogen depletion or dietary induced diuresis (Figure 1).^8^ This work compares the MBM predictions against those derived from the EBT; and it is demonstrated that the MBM not only gives precise descriptions of EBT-consistent data but it also results in remarkably accurate predictions in experimental settings where the EBT makes erroneous forecasts.

**Figure 1.**
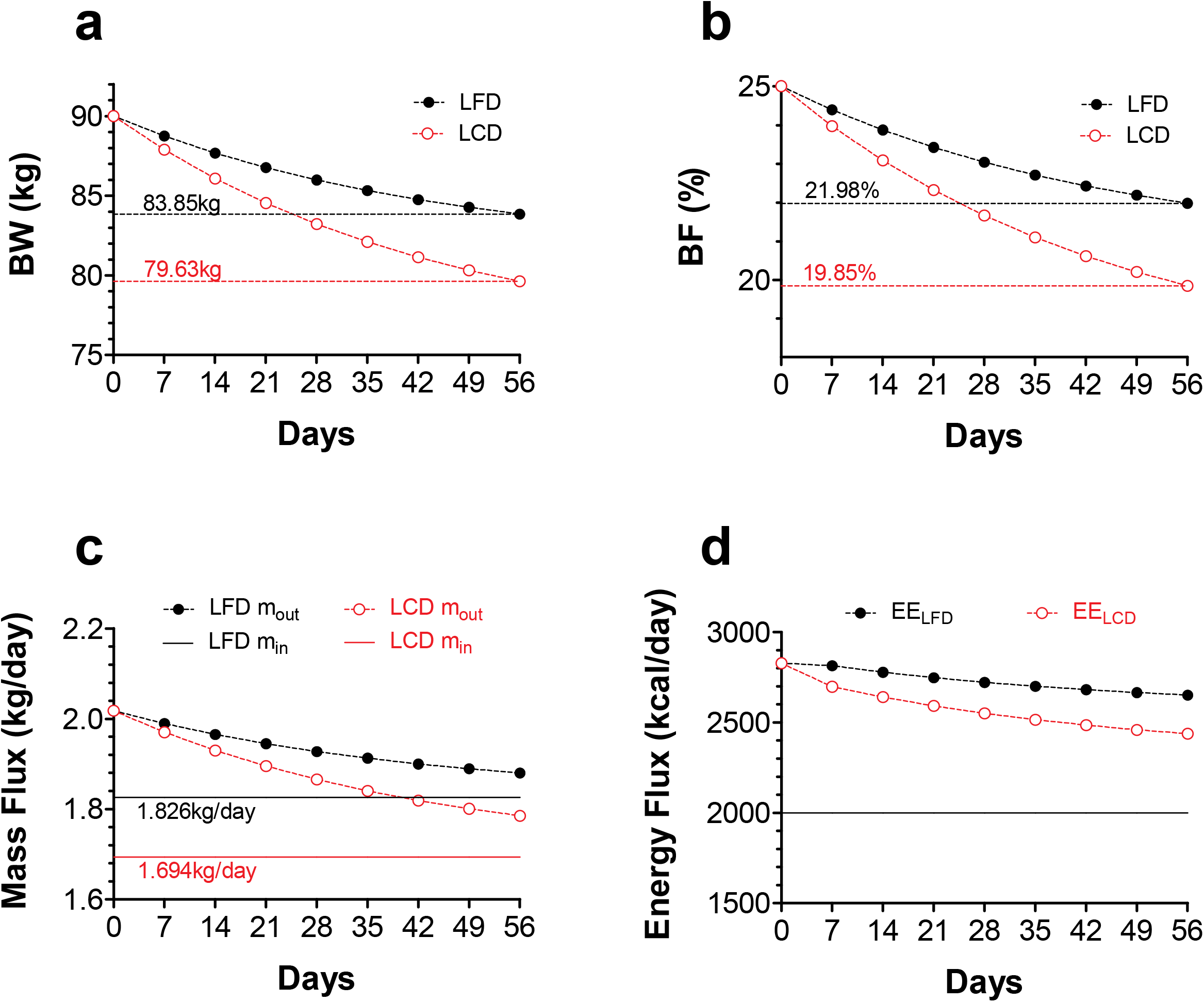
MBM simulation: Energy imbalance magnitude does not predict weight and fat loss outcomes. **a**. Two hypothetical overweight 90kg subjects with identical body composition and weight maintenance energy intakes (EI=2 750 kcal/day; 35% fat (F), 50% carbohydrate (C), 15% protein (P)) initiate two distinct isocaloric diets (2 000 kcal/day): low-fat diet (LFD; 20% F, 65% C, 15% P) vs. low-carbohydrate diet (LCD; 70% F, 15% C, 15% P). The MBM predicts that the LCD results in greater weight loss in contrast to the LFD. **b**. The figure depicts the body fat (BF) percentage changes that correspond to panel a. **C**. As shown, the LCD mass intake (m_in_) is small relative to the eliminated mass (m_out_) and so the net daily mass loss is large (i.e., |m_in_ – m_out_|). In the LFD the net daily mass loss is not as efficient since m_in_ cancels out a substantial fraction of m_out_ decelerating weight loss. LCDs are thus more effective in maximizing the net daily mass loss relative to LFDs and consequently the former manifest a substantially larger cumulative weight loss than the latter. **d**. Daily energy expenditure (EE) data from panel a. The horizontal line represents the daily energy intake. According to the EBT, this graph suggests that the subject in the LFD should lose more weight than the one in the LCD, which is not the case as illustrated in panel a. Energy imbalance is, therefore, not predicting the weight loss outcome in this intervention.

## Materials and Methods

This study compares the MBM predictions against those derived from the EBT. The mathematical structure of each model is described below.

### Mass Balance Model (MBM)

For model derivation and details consult Arencibia-Albite^8^ but, briefly, daily body weight (BW, in kg) fluctuations are given by:

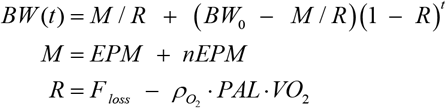

where *t* is time in days; M is the average daily mass intake defined as the sum of the energy-providing mass (EPM; e.g., F, C and P) plus the non-energy-providing mass (nEPM; i.e., water, insoluble fiber, vitamins and minerals); R is the average relative daily rate of mass excretion free of total daily O_2_ uptake; BW_0_ is the initial body weight; F_loss_ is the average relative daily rate of mass excretion that includes of total daily O_2_ uptake; 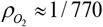 is the average O_2_ density in kg/L; PAL is a dimensionless number that represents the physical activity level computed as the ratio of total daily O_2_ uptake over the total daily resting O_2_ consumption; and VO_2_ is the specific daily resting O_2_ uptake in L/(kg x day).

The EMP parameter obeys the next expression

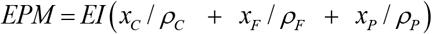

where EI is the energy intake; 0 ≤ *x*_*i*_ ≤ 1 is the energy fraction from *i* = C, F, P; and *ρ*_i_ is the energy density of *i* with *ρ*_*C*_ = 4.2 *kcal* / *g, ρ*_*F*_ = 9.4 *kcal* / *g, ρ*_*C*_ = 4.7 *kcal* / *g*. To simulate pancreatic *β*-cell death *xC* = 0 since the bulk of dietary glucose is lost through urination, i.e., glycosuria. The rational for this is that in untreated type-1 diabetes the characteristic elevated blood glucose concentration saturates the kidney’s monosaccharide reuptake system leading to the appearance of very high quantities of sugar in the urine.

Fat mass (FM) and fat-free mass (FFM) alterations are given by

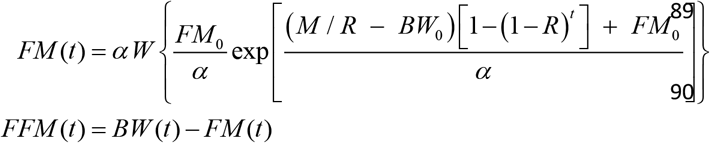

where FM_0_ is the initial fat mass, *α* is a proportionality constant that sets the rate of the change in fat free mass (FFM) whit respect to FM changes, (i.e., dFFM/dFM = *α*/FM) and W is the product log function.

In MBM simulations daily energy expenditure (EE) was estimated by

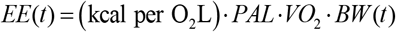

The first term at the right-hand side can be approximated with the diet’s macronutrient composition and the following Weir formula^14^

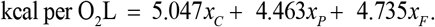

The values of model parameters and algorithms used in simulations are listed below.

Model parameters in Figure 1: BW_0_ = 90kg, α = 10.4kg, FM_0_ = 22.5kg, nEPM = 1.41kg, F_loss_= 0.030833, PAL = 1.5, VO_2_ = 4.32 L/[kg x day].

Model parameters in Figure 2: BW_0_ = 90kg, α = 11kg (HC), α = 10kg (HF), FM_0_ = 19.8kg, nEPM = 1.500kg, F_loss_= 0.030833, PAL = 1.5, VO_2_= 4.32 L/[kg x day].

**Figure 2.**
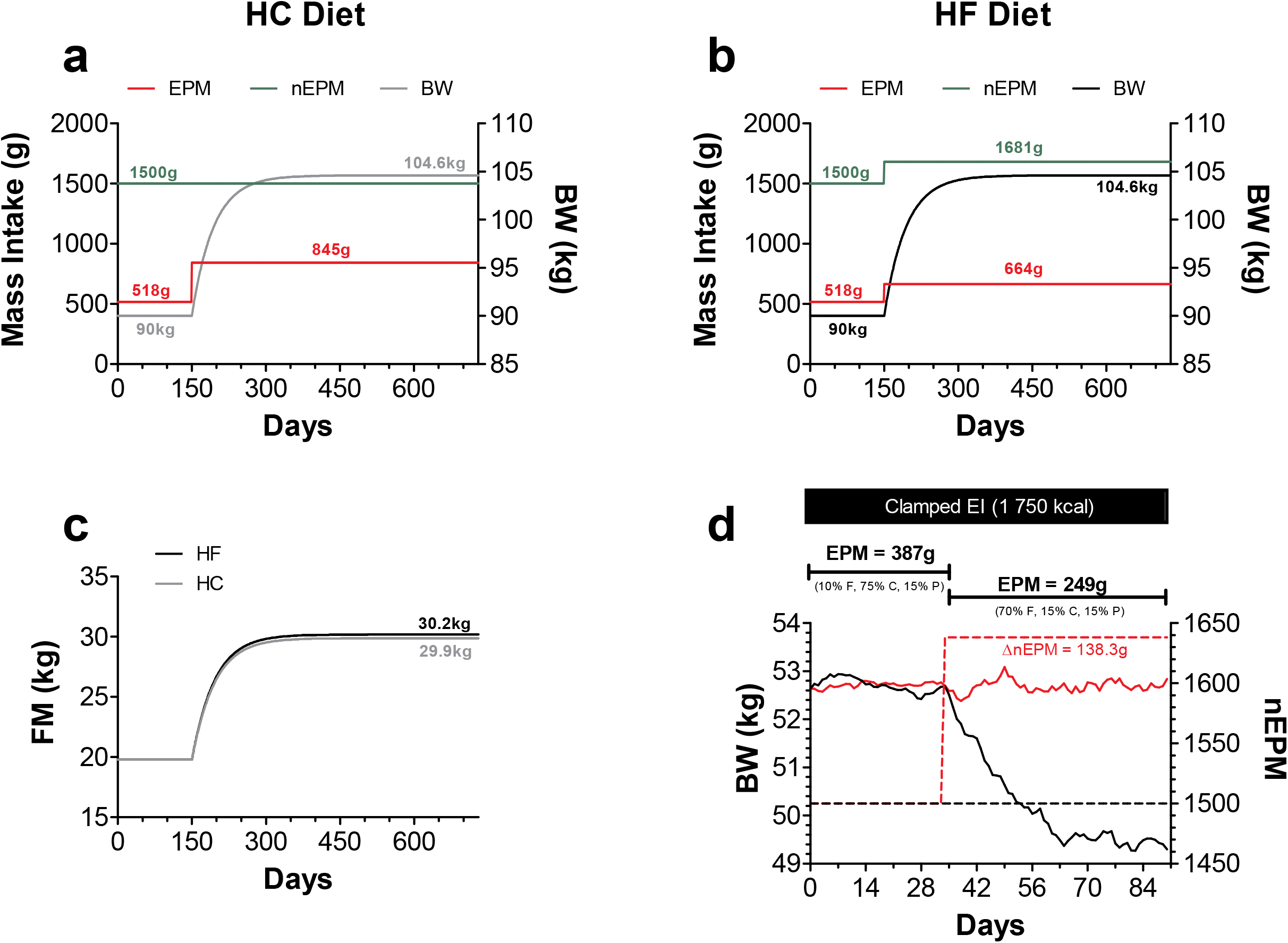
The MBM explains EBT-consistent data. A hypothetical 90kg subject remains weight-stable for 150days (EI=2 750 kcal/day; 35% F, 50%C, 15% P). After day 150 EI increases by 50% (EI=4 125 kcal/day) leading to weight gain. Here excess energy originates only from carbohydrates. The energy-providing mass (EMP) augmented by 327g but the non-energy-providing mass (nEMP) remain fixed at 1 500g. HC: high carbohydrate diet **b**. The same as in a, but excess energy originates only from fat. EPM and nEMP increased by 146g and 181g, respectively. Weight gain trajectory is, however, the same as in a. HF: high fat diet **c**. Fat mass gains corresponding to body weight trajectories in panels a and **d**. Random number simulation that recreates Figure 1 in Leibel *et al*.^18^. A 64 year old female with initial weight of 52.64kg consumes weight maintenance liquid diet of 1 750 kcal for 90 days (BW = 52.84kg at day 90). Before 34 day 10% of EI comes from fat but afterwards this proportion increases to 70%. As illustrated a138.5g increment in nEMP during the high fat diet is sufficient to preserve weight stability after a substantial drop in EPM (red trace). However, if after switching to the high fat diet the nEMP is unaltered then body weight decreases (black trace; BW = 49.3 kg at day 90).

Simulation algorithm in Figure 2 D: *BW* was updated according to the following recurrence relation

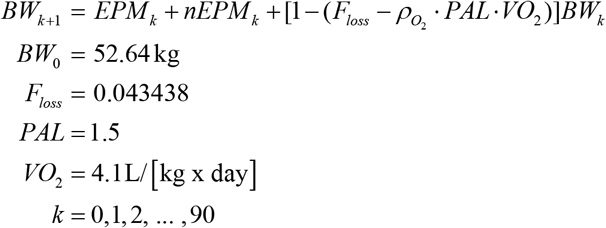

Red trace in Figure 2 D: if *k* < 34 then *EMP*_*k*_ = 0.387 (1 + *CVar* · *x*_*k*_) and *nEMP*_*k*_ = 1.5(1 + *CVar* · *y*_*k*_) otherwise *EMP*_*k*_ = 0.249 (1 *CVar x*_*k*_) and *nEMP*_*k*_ = 1.638(1 + *CVar* · *y*_*k*_), where *CVar* = 0.05 is the coefficient of variation and {*xk, yk*} are random numbers drawn from standard normal distribution.

Black trace in Figure 2 D: if *k* < 34 then *EMP*_*k*_ = 0.387 (1 + *CVar* · *x*_*k*_) and *nEMP*_*k*_ = 1.5(1 + *CVar* · *y*_*k*_) otherwise *EMP*_*k*_ = 0.249 (1 + *CVar y*_*k*_) *CVar* · *x*_*k*_) and *nEMP*_*k*_ = 1.5(1 + *CVar* · *y*_*k*_)

Model parameters in Figure 3: BW_0_ = 70kg, α = 10.4kg, FM_0_ = 14kg, nEPM = 1.5kg, F_loss_ = 0.034942857, PAL = 1.5, VO_2_ = 4.464 L/[kg x day].

**Figure 3.**
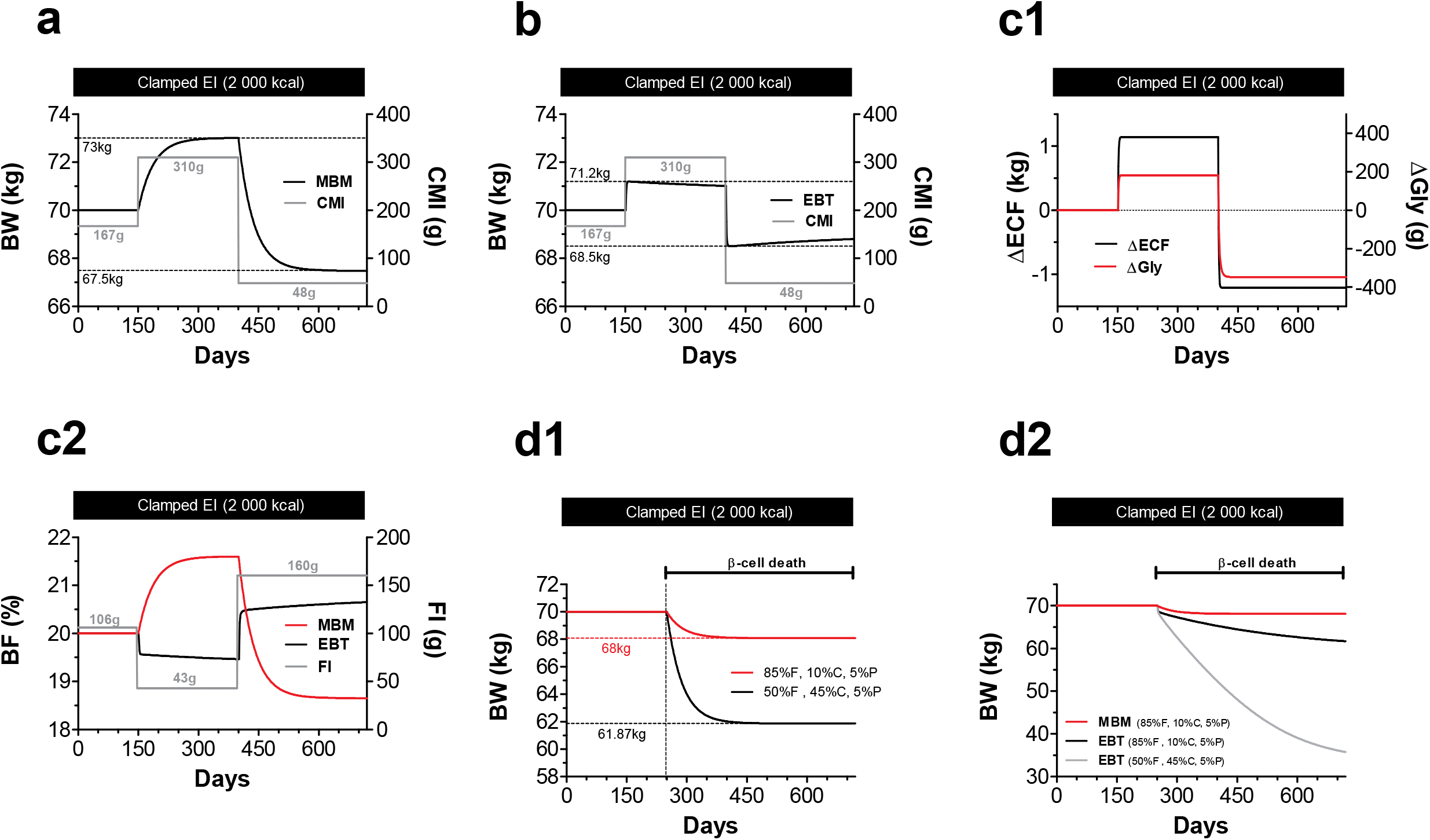
Predictions under isocaloric perturbations: EBT vs. MBM. **a**. MBM response to exchanging a HCD for an isocaloric VLCD. BW is stable at 70 kg with EI = 2 000 kcal during 150 days (control: 50% F, 35% C, 15% P). The daily carbohydrate mass intake (CMI) is 167g. At day 150 EI = 2 000 kcal but CMI has increased by 143g as the control diet is exchange for a HCD (20% F, 65% C, 15% P). On day 400, EI = 2 000 kcal but the diet is exchange again for a VLCD (75% F, 10% C, 15% P). Here CMI decreases by 262g. **b**. EBT response to the same perturbations as in panel a. **c1**. According to the EBT, majority of the weight change under isocaloric perturbations result from changes in extracellular fluid (ECF) and stored glycogen (Gly) that stabilize within few days. **c2**. MBM predicts that BF evolves in parallel to the weight change direction. In contrast, the EBT predicts that, under isocaloric perturbations, BF evolves according to the amount of fat intake (FI). **d1**. MBM predicts that the weight loss magnitude evoked by pancreatic *β*-cell death is dependent on the diet’s macronutrient composition. **d2**. The EBT predicts that changes in the diet’s macronutrient distribution will elicit measurable changes in the rate and magnitude of weight loss after *β*-cell death but not as potent as those predicted by the MBM.

Model parameters in Figure 4: BW_0_ = 65.5kg, α = 10.4kg, FM_0_ = 22.925kg, nEPM_ND_ = 1kg, nEPM_KD_ =0.96kg, F_loss_= 0.029078, PAL = 1.5, VO_2_ = 4.32 L/[kg x day].

**Figure 4.**
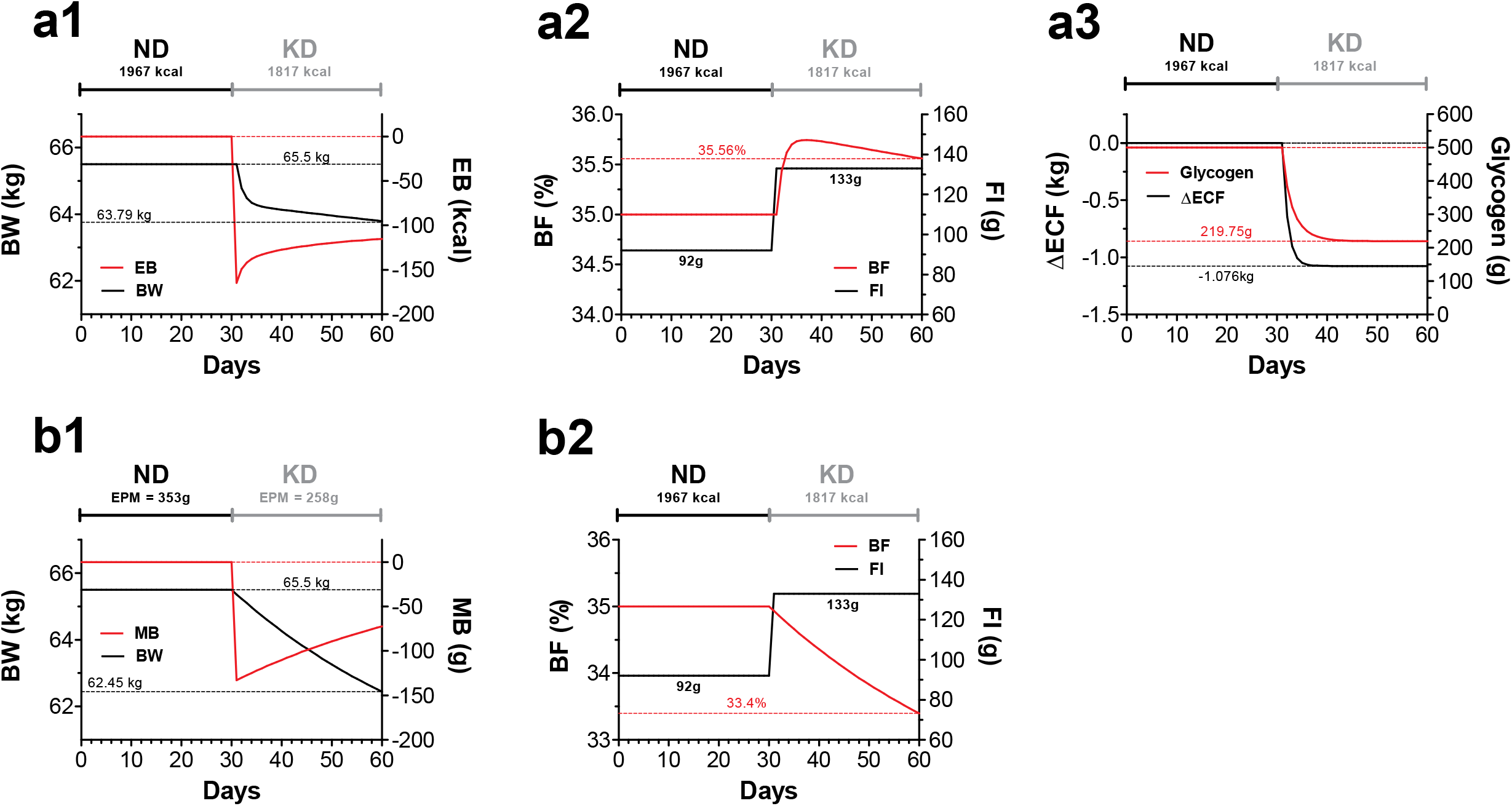
The simulation of Kong *et al*. data: EBT vs. MBM. **a1**. During the normal diet (ND, days 0-30) energy balance (EB, red curve) is zero. After day 30 EB becomes negative under the ketogenic diet (KD) resulting in a 1.71kg weight loss at day 60. **a2**. The EBT predicts that – although the KD results in weight loss – BF increases since FI has augmented by 41g. **a3**. During the ND total glycogen is 500g but after the KD this amount has dropped by 280.25g. ECF has also decreased by 1.076kg. These quantities add to 1.35625kg = 0.28025kg + 1.076kg and, hence, of the 1.71kg weight loss in a1, 0.35375kg = 1.71kg – 1.35625kg are from other mass sources (0.2436 kg fat mass + 0.11015 kg fat-free mass = 0.35375kg). This indicates, according to the EBT, that the total weight loss is distributed as 0.2436 kg of fat plus 1.4664kg of fat-free mass. Consequently, as the decline in fat-free mass is much larger than that of fat mass, BF increases as in a2. **b1**. During the ND period (days 0-30) mass balance (MB, red curve) is zero. After day 30 MB becomes negative under the KD resulting in a 3.05kg weight loss at day 60. **B2**. According to the MBM, of the 3.05kg of weight loss 2.07kg came from fat and 0.98kg from fat-free mass. Even though FI has increased the decline in fat-free mass is much smaller than that of fat mass and thus BF decreases as shown.

*Energy Balance Theory* (EBT): The quantitative form of the EBT use in this study is that of the United States National Institute of Health, NIH, Body Weight Planner developed by Hall *et al*.^15^ A succinct model description is given below.

Glycogen (G), extracellular fluid (ECF), body fat (F_b_), lean tissue (L) and adaptive thermogenesis (AT) are modeled by following system of equations:

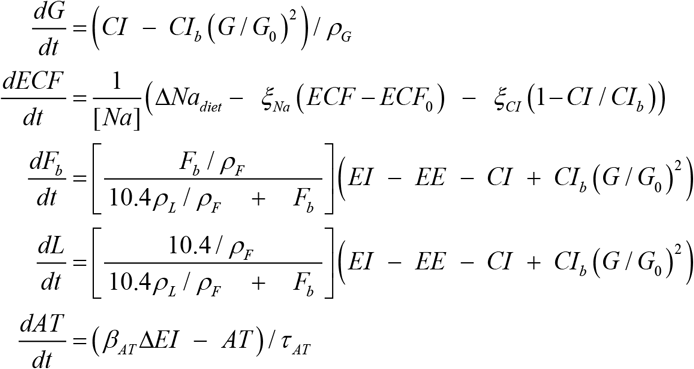

where CI_b_ is the carbohydrate energy intake (CI) at energy balance; G_0_ is the initial body glycogen amount; *ρ*_*G*_ is the glycogen energy density (4.2 kcal/g); [Na] is the extracellular Na concentration (3.22 mg/ml); *ξ*_*Na*_ is the ECF Na excretion constant (3 000 mg/[ml x day]); *ξ*_*CI*_is the CI dependent Na excretion constant (4 000 mg/[ml x day]); *ρ*_*L*_ is the energy density of lean tissue (1.815kcal/g); *τ*_*AT*_ is the AT time constant (14 days); and *β*_*AT*_ = 0.14 is the AT coefficient.

EE is defined as

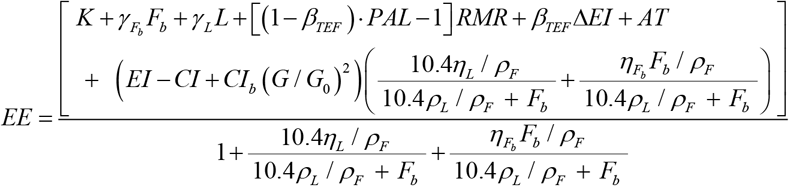

where K is arbitrary constant determined to achieved energy balance at *BW*_0_;

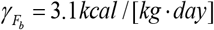 is the specific body fat metabolic rate coefficient; *γ*_*L*_ = 21.99 *kcal* /[*kg*. *day*] is the specific lean tissue metabolic rate coefficient; *β*_*TEF*_ = 0.1 is the thermic effect of feeding coefficient; *RMR* = 19.7(*BW F*) 413 is Mifflin *et al*.^16^ resting metabolic rate formula; *η*_*L*_ = 229.446 *kcal* / *kg* is the lean tissue synthesis efficacy; and 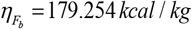 is the fat tissue synthesis efficacy.

Body weight at time *t* is obtained by the following sum

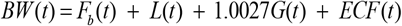

To simulate pancreatic *β*-cell death CI = 0 for the same reasons given in the MBM description. The system of differential equations was solved using the fourth order Runge-Kutta method with a time step of 1 day. The initial conditions are listed below.

Initial conditions for Figure 3: K = 547.2925, BW(0) = 70kg, F_b_(0) = 14kg, L(0) = 36.012kg, ECF(0)=19.486kg, AT(0) = 0, G(0) = 0.5kg, PAL =1.5

Initial conditions for Figure 4: K = 892.7146, BW(0) = 65.5kg, F_b_(0) = 22.925kg, L(0) = 25.698kg, ECF(0) = 16.375kg, AT(0) = 0, G(0) = 0.5kg, PAL =1.5

## Results

### The MBM accounts for EBT-consistent data

In nutrition literature is possible to encounter studies that seem to be in conflict with the MBM’s main hypothesis: body weight fluctuations are a mass imbalance problem and not an energy imbalance issue. Horton *et al*.^17^ overfed, for instance, lean and obese men with isocaloric amounts of either fat or carbohydrate for 2 weeks. Weight gain was similar among treatments even though macronutrient mass intake was greater for carbohydrate overfeeding than for fat overfeeding. Leibel *et al*.^18^, also performed a retrospective analysis of 13 adults and 3 children that lived in metabolic wards and were fed weight-maintenance liquid diets that varied in fat and carbohydrate composition while protein was kept at 15% of the energy requirements. Yet, body weight stability was preserved after switching to an iscocaloric diet of significantly different fat and carbohydrate content. At first glance these EBT-consistent findings may appear to be in disagreement with the MBM; however, as shown next this is not the case.

In the MBM small intake alterations of insoluble fiber, minerals and water are sufficient to account for similar weight gain trajectories or weight stability periods under substantially distinct levels of macronutrient mass ingestion. Figure 2a, 2b and 2c show a situation similar to that reported by Horton *et al*.^17^ were macronutrient mass intake is significantly greater under the high carbohydrate diet (HC) than in the high fat (HF) diet, yet weight and fat gain are similar. To obtain such result the subject in the HF diet has increased the nEPM by 181g (e.g., water intake increases by 181mL). This is a reasonable assumption as increasing fat content in the diet decrease water and minerals present in food which may result in a compensatory fluid and mineral intake. The red trace in Figure 2d recreates Figure 1 in Leibel *et al*.^18^ were decreasing macronutrient mass intake did not alter body weight. Here weight stability was also achieved by a compensatory increment in the nEPM. Nonetheless, if nEPM is not to change after increasing fat content then weight loss will follow (Figure 2d, black trace). Altogether, it shows that the MBM is perfectly capable of explaining data that appears to validate the EBT.

### MBM vs. EBT: effects of macronutrient composition at weight maintenance energy intake

We next performed a theoretical analysis of how changes in carbohydrate and fat content affect body composition when EI is clamped at weight maintenance level. Computational experiments are executed under normal physiology or at the onset of untreated type-1 diabetes (T1D). All the EBT-based simulations were done using the Hall *et al*.^15^ energy balance model.

Figure 3 a uses the MBM to simulate the effect of exchanging a high-carbohydrate diet (HCD) for an isocaloric very-low-carbohydrate diet (VLCD) in a weight-stable 70kg individual. Under the HCD (day 150-400), body weight gradually increases to 73kg. However, after beginning the isocaloric VLCD (day 400), body weight decreases towards a steady value of 67.5kg. Figure 3 b repeats the simulation in panel a, but with the EBT. The latter indicates, in general, that isocaloric perturbations elicit non-significant alterations in body weight that are mostly a consequence of changes in extracellular fluid (ECF) and stored glycogen (Gly, Figure 3 c1). In such cases, therefore, changes in body composition are expected to be nearly undetectable (Figure 3 c2).

We now test the capacity of the MBM and EBT to account for the degree of weight loss observed at the onset of T1D. Before the discovery of insulin, a common dietary treatment of T1D was a very-low-carbohydrate diet/high-fat diet which slowed down and reduced the excessive weight loss while alleviating the classical symptoms of polyuria, polyphagia and polydipsia.^19^ The Figure 3 d1 illustrates that the MBM explains the weight loss associated with the onset of T1D and also makes predictions consistent with the diabetes treatment during the pre-insulin era. The EBT, however, predicts a substantial weight loss at T1D onset even if the pre-onset diet is a VLCD (Figure 3 d2).

Kong *et al*.^6^ have recently published a well-controlled feeding study that allows to test the validity of the predictions made by the MBM and EBT in Figure 3. Their young female subjects (age: 21± standard deviation [SD]: 3.7 years, weight: 65.5 ± SD: 7.7 kg, body mass index: 24.9 ± SD: 2.7 kg/m^2^) were weight-stable for 4 weeks under a normal diet (ND: 1 967kacl ± SD: 362kcal ; F 44% ± SD: 7.6%, C 39.6% ± SD: 5.8%, 15.4%± SD: 3.3%) and then switched to an isocaloric ketogenic diet (KD 1 817kacl ± SD: 285kcal; F 69% ± SD: 5.4%, C 9.2% ± SD: 4.8%, 21.9%± SD: 3.4%) for another 4 weeks leading to a substantially reduced body weight (– 2.9 kg) and body fat percentage (– 2.0%).

Figure 4 a1, a2 and a3 simulate Kong *et al*.^6^ data using the EBT. Here the KD decreases body weight by 1.71kg (Figure 4 a1) with a slight increase in body fat percentage (Figure 4 a2). According to the EBT a substantial fraction of this mass loss is given by reductions in ECF and stored glycogen (Figure 4 a3). Figure 4 b1 and b2 simulate Kong *et al*.^6^ data using the MBM. As illustrated, this model predicts a much larger drop in body weight (–3.05kg, Figure 4 b1) under KD than the EBT. The MBM also predicts a body fat percentage drop of 1.6% (Figure 4 b2).

Therefore, the MBM leads to more accurate predictions than those made by the EBT.

## Discussion

This study demonstrates that when describing distinct dietary interventions with the MBM the main determinant of body composition change is the macronutrient mass intake given that non-substantial differences exists among the remaining model parameters (i.e., nEPM; the average relative daily rate of mass excretion free of total daily O_2_ uptake, R; and the empirical scalar α which sets the rate of change in fat-free mass per change in fat mass). Particularly, if nEPM, R, and α are similar between treatments then the diet that contains the smaller macronutrient mass will lead to greater weight and fat loss during underfeeding (see Figure 1), whereas the diet that has the greater macronutrient mass will result in the largest weight and fat gain during overfeeding. Dissimilar values of the latter parameters, in contrast, may lead to the erroneous impression that MBM is an incorrect model as exemplified in Figure 2. Nonetheless, when these values are taken into consideration, the MBM gives a remarkably accurate description of experimental data that seems to validate the EBT. Therefore, the set of experimental evidence that the MBM is able to explain is apparently much greater than that of the EBT.

According to the EBT, at energy balance, changes in macronutrient composition elicit non-substantial changes in body weight.^20^ These alterations are assumed to be secondary to changes in ECF and glycogen that follow from the diet’s sodium content and carbohydrate intake.^7, 20^ In the EBT body composition changes, under these circumstances, are expected to be small and to occur in parallel to dietary fat intake (see Figure 3 c2). This theory further asserts that blood leptin concentration should be minimally affected as the diet-evoked fat loss or gain is expected to be non-substantial in the aforementioned conditions (see Figure 3 c2). A recent well-controlled feeding trial by Kong *et al*.^6^ showed, however, the opposite. Specifically, they illustrated that the interchange of a weight-preserving normal diet for an isocaloric ketogenic diet lead to a significant decline in body weight, fat mass and leptin levels. From the EBT perspective this finding is a reflection of poor methodology, as body weight and fat alterations are much greater than those predicted when energy balance is assumed to be present (see Figure 4 a1 and a2). This work shows that such EBT-rejected data is, nonetheless, well described and consistent with the MBM (see Figure 4 b1 and b2).

The EBT simulations on pancreatic *β*-cell death, on the other hand, are also inconsistent with a review of case histories from the pre-insulin era illustrating that VLCDs could result in some weight gain after the onset of T1D.^19^ Elliot Proctor Joslin, for example, was the first United States medical doctor that specialized in diabetes treatment during this era. One of his diabetic patients, Mary H., reportedly gained nearly 3kg of body weight while consuming a diet containing solely protein and fat.^19^ The MBM simulations, in contrast to those of the EBT, do predict such therapeutic effect of VLCDs.

Various examples in the weight management literature show that, on average, the amount of weight loss is far greater in LCDs compared to isocaloric LFDs.^1-5^According to the EBT, this can only occur if the energy expenditure under LCDs is larger than that in LFDs. However, in many of these dietary interventions non-significant differences are found between the energy expenditures of these diets (see Figure 1 d).^5,7, 21-23^ The EBT, therefore, argues that in such cases the most likely explanation of the observed superior weight loss is the result of energy intake underreporting by low-fat dieters.^5^ This claim has encountered minimal opposition since it is widely accepted that the majority of the energy intake recording devices (e.g., self-reported food records) are biased toward underestimation.^24, 25^ Two alternative models, however, may explain the apparent weight loss advantage of LCDs over isocaloric LFDs: the carbohydrate-insulin model (CIM) of obesity^26^ and the MBM^8^.

The CIM postulates that high-carbohydrate intake elevates insulin levels leading to the activation of complex neuroendocrine responses that drive body fat deposition, increase appetite and decrease energy expenditure^26^; which, according to the EBT, explains the persistent weight gain observed in obese subjects. Conversely, LCDs, by significantly decreasing circulating insulin levels, should increase energy expenditure by augmenting fat oxidation, which – as argued by CIM advocates – accounts for the greater and faster weight loss observed in feeding trails of LCDs vs. isocaloric LFDs. These claims are open to discussion, however, as evidence shows that obese individuals manifest highly elevated energy expenditures relative to normal weight subjects^27^, plus, as already mentioned, the predominant evidence indicates that no significant energy expenditures differences exist between diets.^5, 7, 21-23^ A recent meta-analysis suggests that an adaptation period of at least 14 days may be required in order for LCDs to elevate energy expenditure.^28^ If this is the case, one would expect a slow decay of the respiratory quotient (RQ) toward 0.71 since this would reflect the gradual metabolic dependence on fat as the main energy fuel. In ketogenic diets, however, RQ reaches a steady state within the first week and continuous to be stable for at least three more weeks.^21^ Taken together, it seems that the LCD-enhanced weight loss is unlikely to be a consequence of augmented energy expenditure as proposed by CIM.

In contrast, the MBM describes body weight fluctuations as a mass imbalance problem; when food and beverage mass intake exceeds the excretion of macronutrient oxidation products body weight increases and *vice versa*. Body weight stability is, hence, expected as over time the average consumed mass equals the average eliminated mass. This model emerges from the following long-standing observations:

### 1. The physiological activity that decreases body weight is the excretion of oxidation products and not energy expenditure^29^

This is exemplified in the oxidation of a general triglyceride molecule:

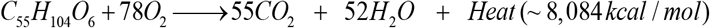

The mass entering this reaction is (in g/mol)

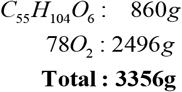

whereas mass exiting the reaction is only present in the reaction products

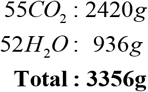

and not in the dissipated heat. Therefore, body mass decreases as the body excretes or eliminates oxidation products (e.g., H_2_O and CO_2_) but not as consequence of the heat in the energy expenditure. Indeed, the oxidation of 1g of fat, glucose or protein will decrease body mass by exactly 1g. Yet, the released heat upon fat oxidation is about twice as large as that glucose or protein oxidation.

### 2. The food property that increases body weight is its mass and not its nutritional energy

For instance, the absorption and retention within body cells of 1 gram of fat, carbohydrate or protein will increase body mass by precisely 1 gram. This observation is independent of the macronutrient kilocalories; according to the Law of Conservation of Mass, the absorbed nutrient mass cannot be destroyed and hence it will contribute to total mass as long as it remains inside the body. Such contribution clearly ends when the macronutrient is eliminated from the body either as oxidation products or in other forms (e.g., shedding of dead skin cells fill with keratin protein).

### 3. Points 1 and 2 imply that body weight fluctuations can be described by the balance between mass intake and mass excretion

Although this statement may seem not to have far-reaching consequences, Arencibia-Albite^8^ has shown that when translated into mathematical form it fits body weight and fat mass data and also results in accurate predictions that are not evident from the qualitative inspection of points 1, 2 and 3. This work further substantiates such predictions and demonstrates that MBM-based simulations result in highly realistic forecasts in settings where the EBT-based simulations collapse.

In today’s energy-centered paradigm of obesity body fat alterations are explained as the balance between fat intake and net fat oxidation. The EBT cannot foresee, therefore, that a LCD may lead to equal or more fat loss than an isocaloric LFD since fat balance in the latter appears to be more negative than in the former.^7^ Plenty of feeding studies suggest, nevertheless, that a LCD can result in equal or greater fat loss than an isocaloric LFD. ^2, 4-6,30^ In a mass-centered framework, these observations are not the result of inadequate methodology; they are, in general, a consequence of the superior negative mass balance evoked by LCD vs isocaloric LFD (e.g., Figure 1 c). From a mass perspective, body fat fluctuations are not only dependent on fat intake and oxidation but other processes contribute to fat loss as well. In a LCD, for instance, the elevated fatty acid oxidation leads, in hepatocytes, to high cytoplasmic HMG-CoA levels which are rapidly reduced to mevalonic acid for cholesterol production. The excess cholesterol of LCDs^31^ is then eliminated in feces as bile acids. Additionally, in LCDs, carbon atoms from the elevated fatty acid breakdown circulate in the blood stream as acetoacetate which can exit the body either in the urine, in sweat^31^ or through breathing when spontaneously decarboxylated to acetone. These processes may be responsible for the often equal or greater fat loss in LCDs vs. isocaloric LFD even if fat oxidation is greater in LFDs as implied by short-term studies.^7^ This suggests, overall, that mass is the fundamental quantity that explains body fat fluctuations and not energy.

In conclusion, the MBM is capable to account for wide range of fending experiments including dietary innervations that seem to agree with the EBT. The EBT, in contrast, is less flexible and results in poor forecasts in settings consistent with the MBM. The widely accepted EBT is thus an unsatisfactory model of body composition fluctuations. Consequently, by shifting to a mass-centered obesity paradigm a much deeper understanding of this terrible disease may follow in the near future.

## Data Availability

N/A.

## Author Contributions

F.A-A. performed experiments; F.A-A. and A.H.M. analyzed and interpreted results of experiments; F.A-A. prepared figures; F.A-A. and A.H.M. drafted manuscript; F.A-A. and A.H.M. edited and revised manuscript; F.A-A. and A.H.M. approved final version of manuscript.

## Data availability

The data that support the findings of this study are available from the corresponding author on reasonable request.

